# Estimating the unseen emergence of COVID-19 in the US

**DOI:** 10.1101/2020.04.06.20053561

**Authors:** Emily M. Javan, Spencer J. Fox, Lauren Ancel Meyers

**Affiliations:** Department of Integrative Biology, University of Texas at Austin, Austin, TX, United States of America; Santa Fe Institute, Santa Fe, New Mexico, United States of America

## Abstract

As SARS-CoV-2 emerged as a global threat in early 2020, China enacted rapid and strict lock-down orders to prevent introductions and suppress transmission. In contrast, the United States federal government did not enact national orders. State and local authorities were left to make rapid decisions based on limited case data and scientific information to protect their communities. To support local decision making in early 2020, we developed a model for estimating the probability of an *unseen* COVID-19 epidemic in each US county based on the number of confirmed cases. We found that counties with only a single reported case by April 13th had a 50% chance that SARS-CoV-2 was already spreading widely. By that date, 85% of US counties covering 96% of the population had reported at least one case. Given the low rates of testing and reporting early in the pandemic, taking action upon the detection of just one or a few cases may be prudent.

**Author Summary:** By March 28, 2020, only 3 months after the first US case of COVID-19 was detected, COVID-19 emerged in all 50 US states. Officials were forced to weigh the economic and societal costs of strict social distancing measures against the future risks of COVID-19 hospitalizations and mortality in their communities. To support local decision makers throughout the US, we developed a simple model to determine the chance that COVID-19 was spreading unseen based on scant reported case counts. In mid-April, 85% of US counties containing 96% of the population had reported at least one confirmed case. Our model predicted that each of those counties thus faced at least a 50% chance that the virus was already spreading widely. Aggressive pandemic mitigation measures, even before a threat is fully apparent, are particularly critical when testing resources are limited.

## Introduction

The pandemic caused by the 2019 novel coronavirus (COVID-19) has claimed over 242,000 American lives as of early November 2020 and may kill tens of thousands more by the end of the year [1-3]. Early in the pandemic, when confirmed case counts were still relatively low across the US, the federal government left decision making largely to state and local public authorities. Amidst great uncertainty, they faced the unprecedented challenge of balancing the threat of a mostly unseen but deadly virus against the economic and societal costs of shelter-in-place and travel restrictions. At the time, most SARS-CoV-2 (the virus responsible for COVID-19) cases were not reported due to the high proportion of mild and asymptomatic infections, limited laboratory testing capacity and strict requirements for receiving tests (e.g. travel or contact with someone from Wuhan, China) [4,5]. The CDC estimated that only one in ten COVID-19 infections were reported during the early phase of the pandemic [6].

As the first cases of COVID-19 were reported, decision makers urgently needed to determine whether these cases reflected sporadic clusters stemming from recent introductions or sustained community transmission that might evolve into a large epidemic. In the southern US, the 2016 expansion of Zika Virus (ZIKV) across the Americas posed a similar challenge. Cryptic transmission meant that by the time a few cases were reported, a large epidemic could already be underway [7]. Here, we describe a stochastic susceptible-exposed-infected-recovered compartmental model framework for estimating the magnitude of an epidemic threat from scarce case data. The approach was originally developed to support situational awareness for ZIKV then adapted for COVID-19. We apply it to estimating the risk of unseen COVID-19 waves in counties across the US during the emergence phase of the pandemic in 2020.

## Results

We modeled the stochastic emergence of COVID-19 accounting for potential superspreading events, asymptomatic infections, and epidemiological characteristics. We assumed all US counties had roughly similar transmission rates. The chance that a county had emerging COVID-19 waves ranged from 9% for zero detected cases to 100% for 25 or more (Fig 1). By March 16, 2020, counties cumulatively reported between 0 and 489 cases totaling 4,009 nationally. Epidemic risk exceeded 50% in roughly 15% of the 3,142 counties covering 63% of the US population. By April 13, 2020, total reported cases in the US climbed to 467,158. Consequently, we estimated that over 85% of US counties comprising 96% of the national population had at least a 50% chance of having an epidemic already underway (Fig 1).

**Fig 1.**
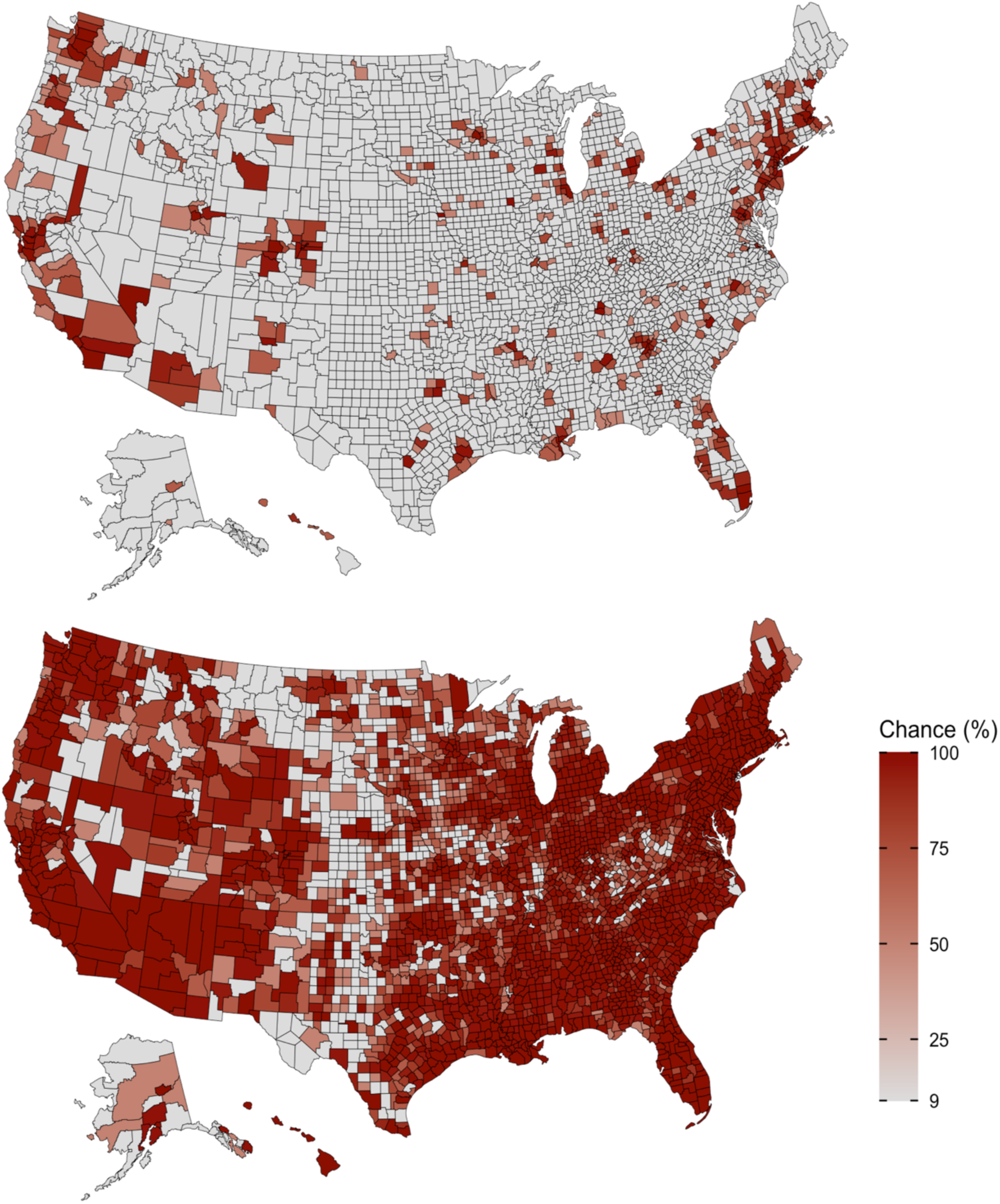
Risk of ongoing COVID-19 epidemics in the 3,142 US counties as of March 16 (top) and April 13, 2020 (bottom). The chance of an unseen epidemic (*epidemic risk*) in a county reporting only a single reported case is 50%; for a county reporting zero cases, the chance is 9%. The model used in both maps assumes *R*_*e*_=1.5, a 10% case detection rate, and a generation time of six days.

Based on COVID-19 case detection rates [6] for the week of April 13, 2020, we estimated that sustained community transmission was probable as soon as even one case was confirmed (Fig 2). At a moderate transmission rate (i.e. *R*_e_=1.5), the first case in a county signals a 50% chance that an epidemic was underway. For a high transmission rate (i.e. *R*_e_=3.0), as may be expected before COVID-19 lockdowns, the estimated risk increased to 83%. The projected risks are generally higher for both larger transmission rates and lowercase detection rates. For example, when *R*_e_ is 1.5, the expected epidemic risk associated with a single case is 50% and increases to 63% when the case detection rate drops from one in ten (10%) to one in twenty (5%). For outbreaks that eventually spread widely, the expected time between the first COVID-19 case report and the epidemic reaching 1,000 cumulative infections was 7.5 (95% CI 3.9-16.3) weeks. The expected time between the tenth reported case and 1,000 cumulative infections shrank by 41% to 4.4 (95% CI 2.1-11.4) weeks (Fig 3).

**Fig 2.**
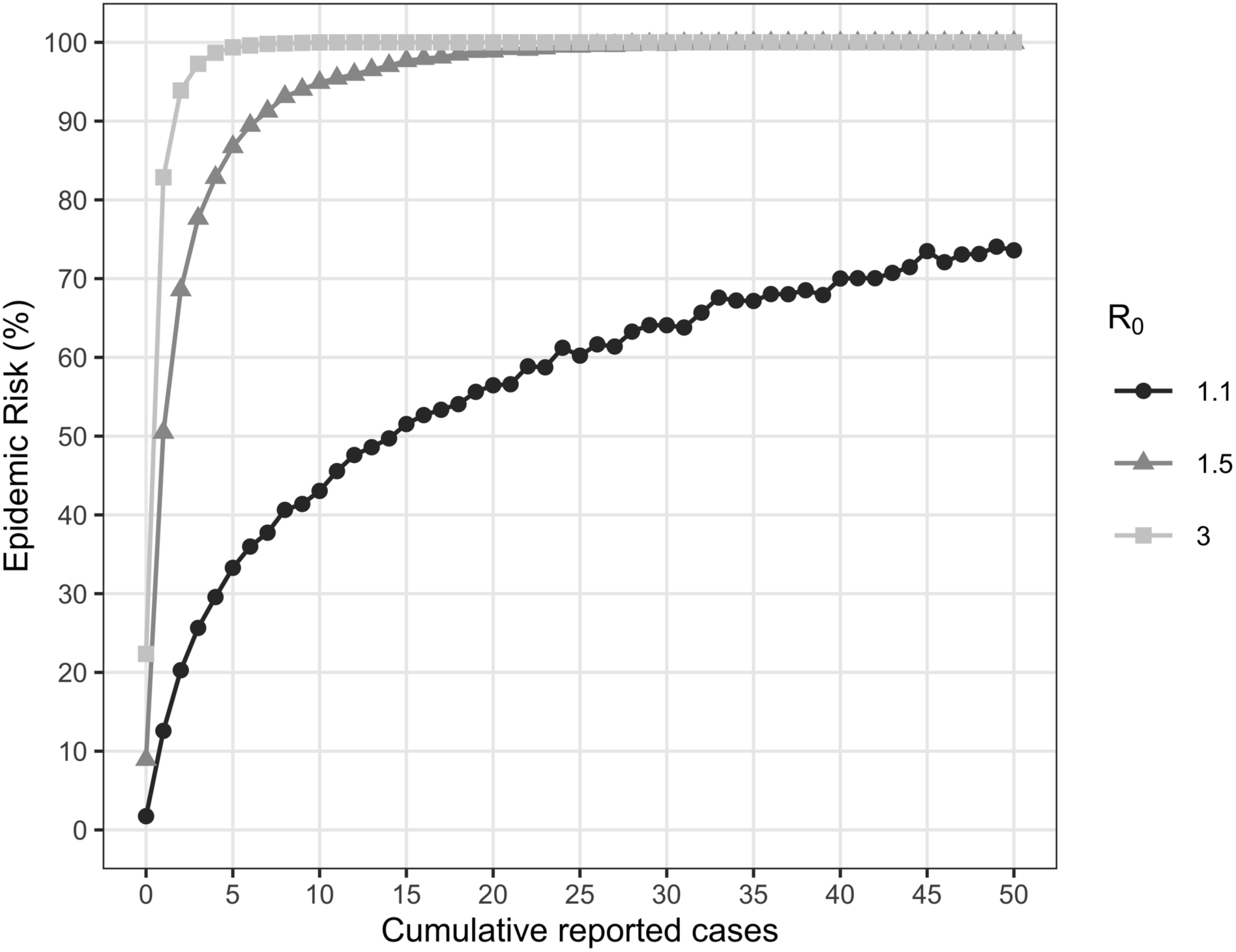
Sensitivity analysis with respect to the effective reproduction number (*R*_e_). For a given number of reported cases, the estimated risk of an epidemic increased with *R*_e_. By the time a single case is reported, there is a 13%, 50% or 83% chance of an ongoing epidemic for *R*_e_ of 1.1, 1.5 or 3.0, respectively.

**Fig 3.**
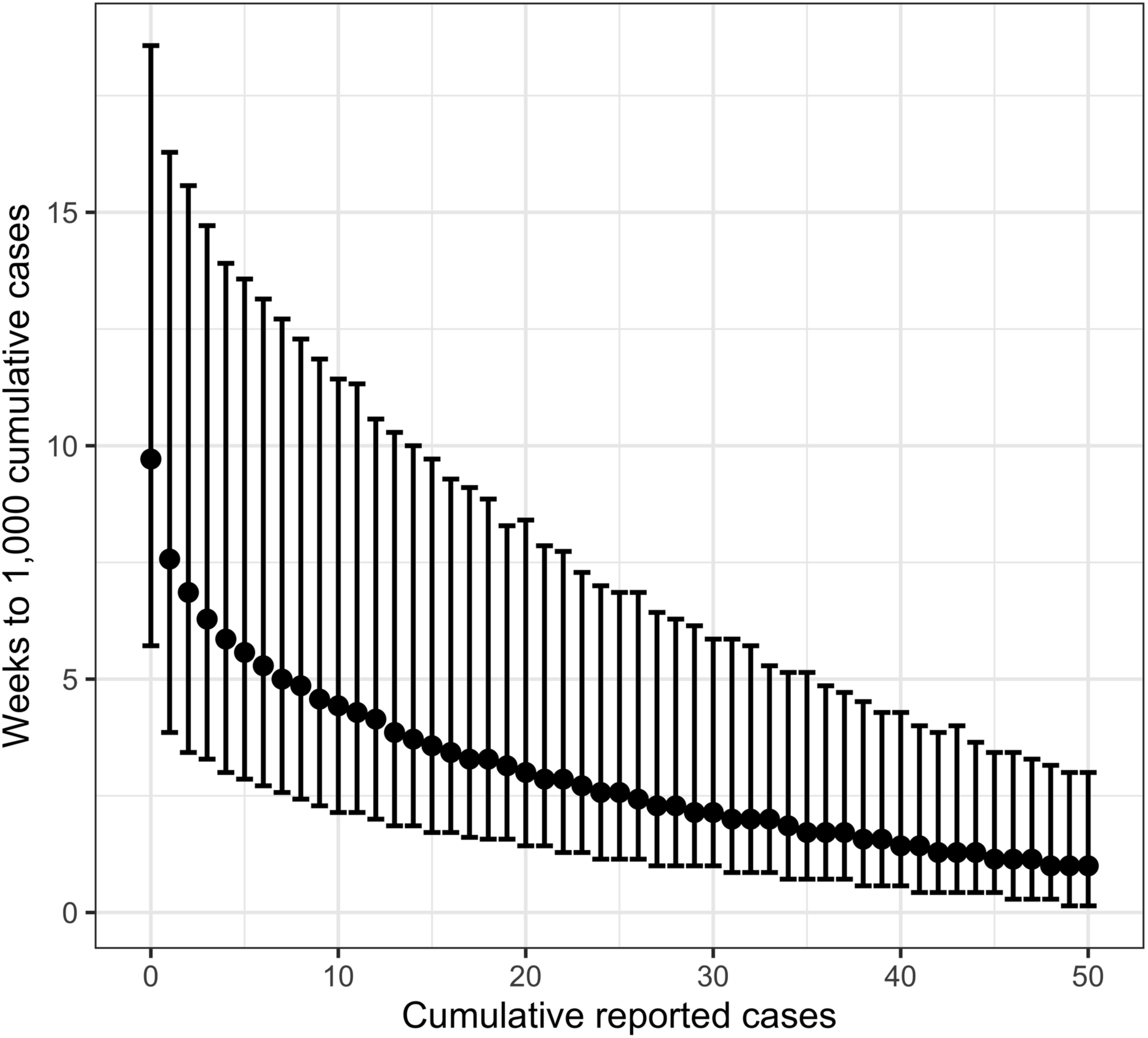
Expected time until the local epidemic exceeds 1,000 cumulative cases. For a given number of cumulative *reported* cases (x-axis), we assume an epidemic is underway then estimate the median and 95% CI (error bars) number of weeks until the cumulative *total* cases exceed 1,000. When the first case is reported, we would expect cumulative cases to surpass 1,000 in 7.5 (95% CI 3.9-16.3) weeks; when the 10th case is reported, the expected lead time shrinks to 4.4 (95% CI 2.1-11.4) weeks. The estimates are based on 100,000 stochastic simulations assuming *R*_*e*_=1.5, a 10% case detection rate, and generation time of six days.

As a retrospective validation of our model, we compared our estimates to reported case counts. We cannot know, with certainty, if and when epidemics began spreading in most US counties. As a proxy, we assess whether case counts increased by at least five in the week following our estimate on March 16 (Fig 4, middle line). We find that our estimates for the probability of an ongoing epidemic (*epidemic risk*) are highly consistent with the fraction of counties that exhibited jumps in reported cases. The cumulative number of reported cases in a county by March 16 was a significant predictor of whether the number of new reported cases in the following week (March 16-23) was at least one, five, or ten cases (logistic regression, *p*<0.001). A one unit increase in cumulative reported cases increased the odds of a county detecting at least one, five, or ten new cases by March 23 by 7.92 (95% CI 5.98-10.80), 4.90 (95% CI 4.14-5.99), and 3.16 (95% CI 2.80-3.63), respectively.

**Fig 4.**
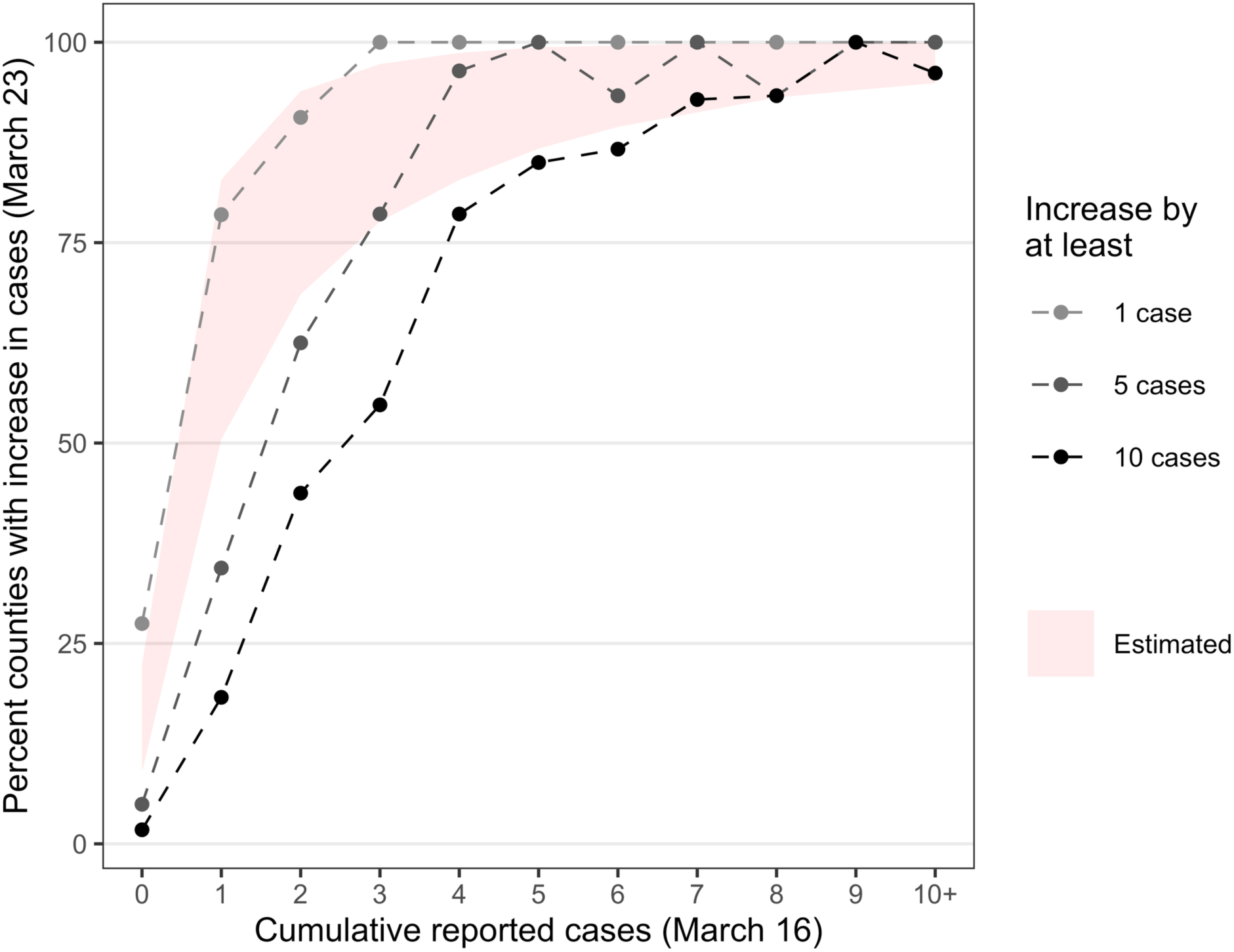
Proportion of all US counties in which COVID-19 case counts increased from March 16 to 23. The light, medium and dark gray lines correspond to increases of at least one, five, or ten new cases within one week, respectively. The red ribbon indicates the model estimates for the probability that an epidemic is underway, given the cumulative reported cases indicated on the x-axis. The bottom and top of the ribbon correspond to scenarios in which *R*_*e*_=1.5 and *R*_*e*_=3.0, respectively. These estimates are calculated based on 100,000 simulations for each reproduction number, assuming a 10% case detection rate and a generation time of six days. The odds of a county detecting at least five new cases increased by 4.90 (95% CI 4.14-5.99) for every one unit increase in cases on March 16. For example, a county with one case on March 16 was roughly five times more likely to have at least six cases a week later than a county with no reported cases.

## Discussion

The timing and rate of COVID-19 emergence varied widely across the US. The earliest of the 3,142 US counties to report a case was Snohomish, Washington on January 21, 2020. By the first of March, April and May, 1%, 70% and 90% of all counties had reported at least one case, respectively. We estimate that, by the time a county reported its first case, it had at least a 50% chance of harboring an unseen but growing epidemic. As of April 13, 2020, the risk exceeded 90% in 54% of counties containing 91% of the US population. The New York Times published real-time projections of our model in a national risk map on April 3, 2020, which spread awareness of the growing COVID-19 threat to the nation [8].

Proactive responses to COVID-19 have been estimated to shorten the duration of costly measures [9,10], whereas delays have likely cost lives [11]. If the goal of COVID-19 interventions is to fully contain an emerging outbreak as quickly as possible, our study suggests that the first reported case should trigger action. The risk of an ongoing epidemic may already exceed 50% and delaying until ten cases have been reported, for example, may substantially reduce the window for corrective action and amassing adequate healthcare and other mitigation resources.

Our analyses make several key assumptions. Case detection rates may vary geographically and change through time depending on testing availability and regulations. Our assumption of 10% is based on a CDC seroprevalence study, which reported rates ranging from 4% to 16% across ten sites [6]. We modeled superspreading events based on estimates for SARS-CoV in Singapore in 2003 [12], which are consistent with more recent reports for SARS-CoV-2 [13–15]. Our estimates do not account for repeated importations given the stay at home orders and travel restrictions at the time. Multiple introductions would reduce our estimated levels of epidemic risk since detected cases could reflect independent clusters rather than continuous chains of transmission. Finally, our estimates depend on the effective reproduction number of the pandemic which can vary spatiotemporally depending on local policies, testing efforts, behavior, and population density [16,17]. Our estimates for mid-April, when much of the US was under shelter-in-place orders, assume a relatively low *R*_*e*_ of 1.5. Our retrospective validation using data from mid-March, when intervention efforts varied geographically, considers reproduction numbers ranging from 1.5 to 3.0.

This analysis, while simple, provided useful insight during a highly uncertain period of the COVID-19 pandemic and can be easily adapted to provide early situational awareness for future emerging infectious outbreaks. Our results suggest that proactive control measures may be prudent, even before the threat becomes apparent [18].

## Methods

We obtained county-level estimates for confirmed and suspected COVID-19 cases from a data repository curated by the New York Times [19] and 2019 estimates of each county’s population from the US Census Bureau [20]. We adapted the framework of another silent spreader–Zika Virus (ZIKV)–which threatened to emerge in southern US states in 2016 [7] to model COVID-19 in US counties. The discrete-time SEIR model assumes a branching process for early transmission in which the number of secondary infections per infected case is distributed according to a negative binomial distribution to capture occasional superspreading events, as estimated for SARS-CoV outbreaks in 2003 [12]. The exposure and infectious periods consist of “boxcars” to enforce the minimum number of days simulated individuals spent in each compartment. We account for imperfect detection and COVID-19 specific epidemiological characteristics (details in Table 1). Our baseline scenario assumes the *R*_*e*_ of COVID-19 is 1.5, accounting for ongoing social distancing measures across the US by mid-April, 2020 [21], and 10% detection of all cases. We do not explicitly model asymptomatic or pre-symptomatic transmission and thus maintain a low detection probability for all infectious cases. To assess the impact of these assumptions on our estimates, we conducted a sensitivity analysis that varied *R*_*e*_ (1.1 and 3.0) and detection rates (5%-40%).

**Table 1.** Model parameters used for simulating COVID-19 outbreaks.

| Parameter | Description | Estimate | Source |
| --- | --- | --- | --- |
| $R_e$ | Effective reproduction number: Average number of new cases from one infected individual in a susceptible and non-susceptible population | 1.5<br>1.1, 3.0 | [22]<br>[17] |
| $T_G$ | Generation time (days): Average length of time between consecutive exposures<br>$T_G = \frac{e}{\nu} + \left(\frac{1}{2}\right)\frac{n}{\delta} = T_E + \left(\frac{1}{2}\right)T_I$ | 6 | [23,24] |
| $T_E$ | Latent period (days) | 1.25 | Fit to $T_G$ |
| $T_I$ | Infectious period (days) | 9.5 | [23] |
| $e$ | Number of exposed compartments in boxcar implementation (min days of exposure) | 1 | Fit to $T_G$ |
| $n$ | Number of infectious compartments in boxcar implementation (min days of infectiousness) | 7 | [23] |
| $\nu$ | Incubation rate: Daily probability of progressing from one exposed compartment to the next | 0.80 | Fit to $T_G$ |
| $\delta$ | Recovery rate: Daily probability of progressing from one infectious compartment to the next | 0.73 | Fit to $T_I$ |
| $\eta$ | Daily detecting rate: The daily probability of an infectious individual being detected, $\frac{0.1}{T_I}$ | 0.01 | [25] |
| $k$ | Total dispersion parameter of negative binomial distribution | 0.16 | [12] |
| | R code for number of new infectious individuals drawn daily:<br>$rbinom(n = 1, prob = \frac{k}{R_0 + k}, size = \frac{k}{T_I})$ | | |

We ran 100,000 stochastic outbreak simulations per scenario beginning with a single undetected case and ending when cumulative infections reached 2,000 or the outbreak died out (whichever came first). Because we model transmission as a branching process, the susceptible population does not deplete as in other compartmental SEIR models. Following the methodology of [7], simulated outbreaks that reached 2,000 cumulative cases and had a minimum prevalence of 50 cases per day were classified as epidemics. We calculated the probability of an epidemic for a given number of detected cases, *x*, by looking at all outbreaks that had *x* reported cases and calculating the proportion of those outbreaks that progressed to epidemics. We then matched county case numbers with the detected case number to obtain epidemic probabilities for each US county based on their reported cumulative number of cases. We use our baseline scenario to compare US maps of epidemic risk from March 16 and April 13, although *R*_*e*_closer to 3.0 may be appropriate for mid-March. For simulations that became epidemics, we also calculated the distribution of lags (in weeks) between the day the *x*th case was reported and the day the epidemic surpassed 1,000 cumulative cases. Confidence intervals were calculated with the *quantile* function in R version 3.6.1 [26].

To validate epidemic risk, we fit three logistic regressions to if US counties reported at least one, five, or ten new cases over the week of March 16 to 23, 2020 (y-axis) and how many cumulative cases there were on March 16 (x-axis). First, counties were grouped by the number of reported cases on March 16. Counties with ten or more cases were put into one group due to the low number of counties with more than ten cases on March 16. Second, March 23rd case counts were subtracted from those on March 16 and the difference was classified as an increase of at least one, five, or ten cases (three separate binary classifications). Finally, a logistic regression was fit to each classification to determine if the number of cases on March 16 was a significant predictor of new cases one week later. This week in mid-March was before lockdowns took place in the US and saw only a moderate increase in daily tests nationally (from 20,000 to 60,000) [27]. We compare case counts from Monday to Monday to avoid weekend reporting bias. To readily compare with epidemic risk estimates, we plot the percent of counties with an increase in new cases with our epidemic risk estimates from *R*_*e*_=1.5 to *R*_*e*_=3.0.

## Data Availability

Code to generate data available online.

https://github.com/MeyersLabUTexas/COVID-19-Epidemic-Risk

https://github.com/sjfox/rtZIKVrisk

## References

1. Woody S, Tec MG, Dahan M, Gaither K, Lachmann M, Fox SJ, et al. Projections for first-wave COVID-19 deaths across the US using social-distancing measures derived from mobile phones. medRxiv:2020.04.16.20068163v2 [Preprint]. 2020 [cited 2020 Nov 11]. Available from: https://www.medrxiv.org/content/10.1101/2020.04.16.20068163v2

2. IHME COVID-19 health service utilization forecasting team, Murray CJL. Forecasting COVID-19 impact on hospital bed-days, ICU-days, ventilator-days and deaths by US state in the next 4 months. medRxiv:2020.03.27.20043752v1 [Preprint]. 2020 [cited 2020 Nov 11]. Available from: https://www.medrxiv.org/content/10.1101/2020.03.27.20043752v1

3. Centers for Disease Control and Prevention. COVID-19 forecasts: deaths. 2020 [cited 2020 Nov 5]. Database: CDC [Internet]. Available from: https://www.cdc.gov/coronavirus/2019-ncov/covid-data/forecasting-us.html

4. Li R, Pei S, Chen B, Song Y, Zhang T, Yang W, et al. Substantial undocumented infection facilitates the rapid dissemination of novel coronavirus (SARS-CoV-2). Science. 2020;368: 489–493. doi: 10.1126/science.abb3221.

5. Ansari FM, Aggarwal K, Chopra A, Agrawal MG, Soni P, Agarwal P, et al. Asymptomatic coronavirus: A Boon or Bane?. JAMDSR. 2020;8: 109–111. doi: 10.21276/jamdsr.

6. Havers FP, Reed C, Lim T, Montgomery JM, Klena JD, Hall AJ, et al. Seroprevalence of Antibodies to SARS-CoV-2 in 10 Sites in the United States, March 23-May 12, 2020. JAMA Intern Med. 2020. doi: 10.1001/jamainternmed.2020.4130.

7. Castro LA, Fox SJ, Chen X, Liu K, Bellan SE, Dimitrov NB, et al. Assessing real-time Zika risk in the United States. BMC Infect Dis. 2017;17: 1–9. doi: 10.1186/s12879-017-2394-9.

8. Glanz J, Bloch M, Singhvi A. Does my county have an epidemic? Estimates show hidden transmission. The New York Times. 2020 Apr 4 [cited 2020 Nov 11]. Available from: https://www.nytimes.com/interactive/2020/04/03/us/coronavirus-county-epidemics.html

9. Du Z, Xu X, Wang L, Fox SJ, Cowling BJ, Galvani AP, et al. Effects of proactive social distancing on COVID-19 outbreaks in 58 cities, China. Emerg Infect Dis. 2020;26: 2267–2269. doi: 10.3201/eid2609.201932.

10. Lyu W, Wehby GL. Community use of face masks and COVID-19: evidence from a natural experiment of state mandates in the US. Health Aff. 2020;39: 1419–1425. doi: 10.1377/hlthaff.2020.00818.

11. Pei S, Kandula S, Shaman J. Differential effects of intervention timing on COVID-19 spread in the United States. medRxiv:2020.05.15.20103655v2 [Preprint]. 2020 [cited Nov 11 2020]. Available from: https://www.medrxiv.org/content/10.1101/2020.05.15.20103655v2

12. Lloyd-Smith JO, Schreiber SJ, Kopp PE, Getz WM. Superspreading and the effect of individual variation on disease emergence. Nature. 2005;438: 355–359. doi: 10.1038/nature04153.

13. Adam D, Wu P, Wong J, Lau E, Tsang T, Cauchemez S, et al. Clustering and superspreading potential of SARS-CoV-2 infections in Hong Kong. Nat Med. 2020. doi: 10.1038/s41591-020-1092-0.

14. Zhang Y, Li Y, Wang L, Li M, Zhou X. Evaluating transmission heterogeneity and super-spreading event of COVID-19 in a metropolis of China. Int J Environ Res Public Health. 2020;17: 3705. doi:10.3390/ijerph17103705.

15. Endo A, Centre for the Mathematical Modelling of Infectious Diseases COVID-19 Working Group, Abbott S, Kucharski AJ, Funk S. Estimating the overdispersion in COVID-19 transmission using outbreak sizes outside China. Wellcome Open Res. 2020;5: 67. doi: 10.12688/wellcomeopenres.15842.3.

16. Ives AR, Bozzuto C. Estimating and explaining the spread of COVID-19 at the county level in the USA. medRxiv:2020.06.18.20134700v4 [Preprint]. 2020 [cited 2020 Nov 11]. Available from: https://www.medrxiv.org/content/10.1101/2020.06.18.20134700v4

17. Sy KTL, White LF, Nichols BE. Population density and basic reproductive number of COVID-19 across United States counties. medRxiv:2020.06.12.20130021v1 [Preprint]. 2020 [cited 2020 Nov 11]. Available from: https://www.medrxiv.org/content/10.1101/2020.06.12.20130021v1

18. Cowling BJ, Ali ST, Ng TWY, Tsang TK, Li JCM, Fong MW, et al. Impact assessment of non-pharmaceutical interventions against coronavirus disease 2019 and influenza in Hong Kong: an observational study. Lancet Public Health. 2020;5: e279–e288. doi: 10.1016/S2468-2667(20)30090-6.

19. The New York Times. Coronavirus (Covid-19) data in the United States; 2020 [cited 2020 Nov 11]. Database: github [Internet]. Available from: https://github.com/nytimes/covid-19-data

20. US Census Bureau. County population totals: 2010-2019; 2020 [cited 2020 Nov 11]. Database: census [Internet]. Available from: https://www.census.gov/data/tables/time-series/demo/popest/2010s-counties-total.html

21. Koo JR, Cook AR, Park M, Sun Y, Sun H, Lim JT, et al. Interventions to mitigate early spread of SARS-CoV-2 in Singapore: a modelling study. Lancet Infect Dis. 2020;20: 678– 688. doi: 10.1016/S1473-3099(20)30162-6.

22. Shim E, Tariq A, Choi W, Lee Y, Chowell G. Transmission potential and severity of COVID-19 in South Korea. Int J Infect Dis. 2020;93: 339–344. doi: 10.1016/j.ijid.2020.03.031.

23. He X, Lau EHY, Wu P, Deng X, Wang J, Hao X, et al. Temporal dynamics in viral shedding and transmissibility of COVID-19. Nat Med. 2020;26: 672–675. doi: 10.1038/s41591-020-0869-5.

24. Bi Q, Wu Y, Mei S, Ye C, Zou X, Zhang Z, et al. Epidemiology and transmission of COVID-19 in 391 cases and 1286 of their close contacts in Shenzhen, China: a retrospective cohort study. Lancet Infect Dis. 2020;20: 911–919. doi: 10.1016/S1473-3099(20)30287-5.

25. Perkins A, Cavany SM, Moore SM, Oidtman RJ, Lerch A, Poterek M. Estimating unobserved SARS-CoV-2 infections in the United States. PNAS. 2020;117: 22597–22602. doi: 10.1073/pnas.2005476117.

26. R Core Team. R: a language and environment for statistical computing. Version 3.6.1 [software]. 2019. Available: https://www.R-project.org/

27. The Atlantic. The COVID tracking project US historical data; 2020 [cited 2020 Nov 11]. Database: covidtracking [Internet]. Available from: https://covidtracking.com/data/national

